# Cryptic transmission of SARS-CoV-2 in Washington State

**DOI:** 10.1101/2020.04.02.20051417

**Authors:** Trevor Bedford, Alexander L. Greninger, Pavitra Roychoudhury, Lea M. Starita, Michael Famulare, Meei-Li Huang, Arun Nalla, Gregory Pepper, Adam Reinhardt, Hong Xie, Lasata Shrestha, Truong N Nguyen, Amanda Adler, Elisabeth Brandstetter, Shari Cho, Danielle Giroux, Peter D. Han, Kairsten Fay, Chris D. Frazar, Misja Ilcisin, Kirsten Lacombe, Jover Lee, Anahita Kiavand, Matthew Richardson, Thomas R. Sibley, Melissa Truong, Caitlin R. Wolf, Deborah A. Nickerson, Mark J. Rieder, Janet A. Englund, the Seattle Flu Study Investigators, James Hadfield, Emma B. Hodcroft, John Huddleston, Louise H. Moncla, Nicola F. Müller, Richard A. Neher, Xianding Deng, Wei Gu, Scot Federman, Charles Chiu, Jeff Duchin, Romesh Gautom, Geoff Melly, Brian Hiatt, Philip Dykema, Scott Lindquist, Krista Queen, Ying Tao, Anna Uehara, Suxiang Tong, Duncan MacCannell, Gregory L. Armstrong, Geoffrey S. Baird, Helen Y. Chu, Jay Shendure, Keith R. Jerome

## Abstract

Following its emergence in Wuhan, China, in late November or early December 2019, the SARS-CoV-2 virus has rapidly spread throughout the world. Genome sequencing of SARS-CoV-2 strains allows for the reconstruction of transmission history connecting these infections. Here, we analyze 346 SARS-CoV-2 genomes from samples collected between 20 February and 15 March 2020 from infected patients in Washington State, USA. We found that the large majority of SARS-CoV-2 infections sampled during this time frame appeared to have derived from a single introduction event into the state in late January or early February 2020 and subsequent local spread, indicating cryptic spread of COVID-19 before active community surveillance was implemented. We estimate a common ancestor of this outbreak clade as occurring between 18 January and 9 February 2020. From genomic data, we estimate an exponential doubling between 2.4 and 5.1 days. These results highlight the need for large-scale community surveillance for SARS-CoV-2 and the power of pathogen genomics to inform epidemiological understanding.

## Main text

The novel coronavirus, referred to alternately as SARS-CoV-2 *(2)* or hCoV-19 *(3)*, emerged in Wuhan, Hubei, China, in late November or early December 2019 *(4)*. As of 25 March 2020, COVID-19, the disease caused by SARS-CoV-2, COVID-19, has globally caused 413,467 confirmed cases and 18,433 deaths *(5)*. After its initial emergence in China, travel-associated cases with travel histories related to Wuhan appeared in other parts of the world *(6)*. The first confirmed case in the United States was travel-associated and was detected in Snohomish County, Washington State, on 19 January 2020. Until 27 February 2020, the US Centers for Disease Control and Prevention (CDC) guidance recommended focusing testing on persons with direct travel history or exposure to a known case—cases of respiratory disease with no known risk factors were not routinely tested. In the 6 weeks between 19 January and 27 February, 59 confirmed cases were reported in the United States *(7)*, all with either direct travel history or exposure to a known confirmed case. On 28 February 2020, a community case was identified close to the location of the original Snohomish County case *(8)*. By 25 March 2020, in the setting of continuing transmission and increased testing, Washington State had reported 2580 confirmed cases and 132 deaths *(9)*.

Here we report on the putative history of community transmission in Washington State as revealed by genomic epidemiology. We conclude that SARS-CoV-2 was circulating cryptically, ie undetected by the surveillance apparatus, in Washington State since January 2020, and we underscore the following recommendations in settings where large-scale community transmission is not yet recognized: the importance of early identification of the virus, extensive testing of potential cases, and immediate self-isolation of infected persons.

All SARS-CoV-2 genomes represented in convenience samples from the COVID-19 pandemic appear closely genetically related with the large majority possessing between 0 and 12 mutations relative to a common ancestor estimated to exist in Wuhan between late Nov and early December 2019 (**Supplementary Fig. 1**). This pattern is consistent with a reported rate of molecular evolution of ∼0.8 × 10^−3^ substitutions per site per year or ∼2 substitutions per genome per month *(4)*. After its initial emergence by the zoonotic route in Wuhan *(10)*, SARS-CoV-2 viral genomes began to accumulate substitutions and spread from Wuhan to other regions in the world *(4)*. During December 2019, the Wuhan outbreak was too small to seed many introductions outside of China, but by January 2020, it had grown large enough to begin seeding cases elsewhere in the world *(11)*. At this point, travel cases with origins in Hubei began to appear in the US. The first confirmed case recorded in the United States was a travel-associated case from an individual returning from Wuhan on 15 January 15 2020 who presented for care at an outpatient clinic in Snohomish County on 19 January 2020 and tested positive *(12)*. This infection is recorded as strain USA/WA1/2020 (referred to here as WA1) and appears closely related to viruses from infections in China (Fujian, Hangzhou and Guangdong provinces). Relative to the basal virus at root of the phylogeny, WA1 possesses mutations C8782T and T28144C (found in 74/224 sampled viruses from China) alongside C18060T (found in 6/224 sampled viruses from China).

Sequencing of viruses from the Washington State outbreak began on 28 February 2020 and has continued since then. We analyzed the sequences of 346 SARS-CoV-2 viruses from the Washington State outbreak collected between 20 February and 15 March 2020. The large majority (293, 85%) of these viruses fall into a closely related clade, and all share the mutations possessed by WA1 and the additional mutations C17747T and A17858G. This results in the nested tree structure shown in **Figure 1A** in which the Washington State outbreak viruses group together and appear as “direct descendants” of WA1 in a maximum likelihood tree with branches measuring substitutions. This tree structure is consistent with the WA1 strain transmitting locally after arrival into the United States. However, because the rate of evolution of 1 mutation per ∼15 days is slower than the transmission rate calculated for SARS-CoV-2 of 1 transmission event every 4-8 days *(13, 14)*, it is also possible that WA1 sits on a side branch of the underlying transmission tree even if it appears as a direct ancestor in the maximum likelihood tree.

**Figure 1.**
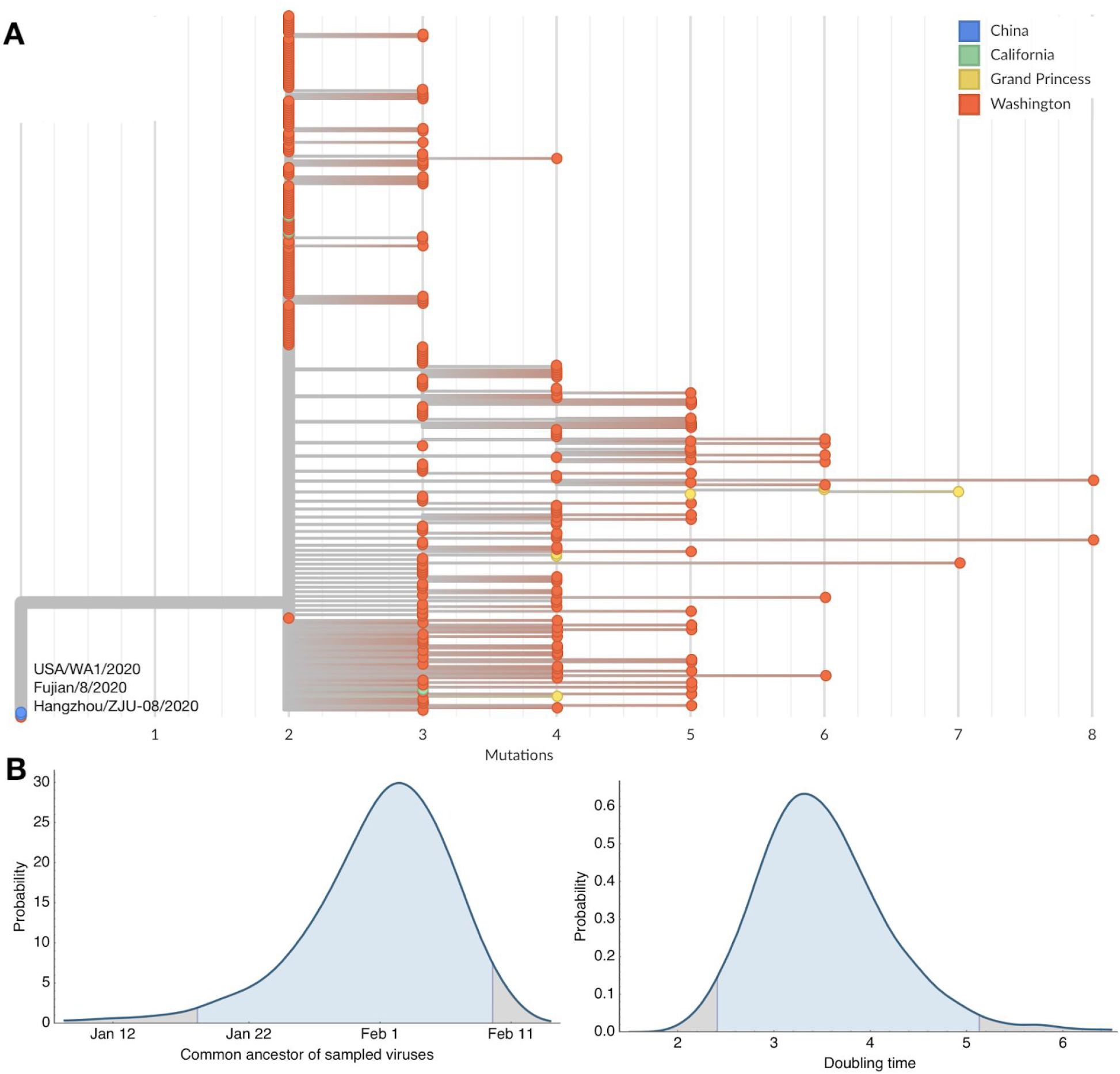
Maximum-likelihood resolved phylogeny of WA1 outbreak clade containing 303 SARS-CoV-2 viruses (A) and Bayesian estimates of outbreak ancestor and doubling time (B). (A) Branch lengths are proportional to the number of substitutions between viruses and the x-axis is labeled with the number of substitutions relative to the root of the overall SARS-CoV-2 phylogeny. Tips on the phylogeny are colored based on location of sampling with viruses from China in blue, viruses from California in green, viruses from the Grand Princess cruise ship in yellow and viruses from Washington State in red. This comb-like phylogenetic structure is consistent with rapid exponential growth of the virus population. (B) Highest posterior density estimates for the date of the common ancestor of viruses from the Washington outbreak clade as well as the doubling time in days of the growth of this clade.

We sought to test these two hypotheses: (a) SARS-CoV-2 was introduced into Washington State on 15 January 2020 with the arrival of WA1; subsequent cryptic transmission led to a community outbreak first detected on 28 February 2020 and (b) SARS-CoV-2 was imported on 15 January 2020 but this infection did not transmit onwards; a second, initially undetected importation event of a genetically identical or highly similar virus occurred, followed by cryptic transmission that led to a community outbreak. The fact that only a small proportion of infections in China have been sequenced makes it difficult to test these two hypotheses. We attempted a simple probability calculation by noting that the C8782T, T28144C, C18060T variant appears in only six (Fujian/8/2020, Chongqing/YC01/2020, Hangzhou/ZJU-08/2020, Guangdong/GD2020086-P0021/2020, Guangdong/GD2020234-P0023/2020, Guangdong/FS-S30-P0052/2020) out of a total of 224 viral genomes from mainland China. If another variant had been introduced to Washington State, it would not result in the observed nesting pattern. Thus, an extremely rough probability calculation is that there is a 6/224 or 3% chance of observing this pattern under hypothesis (b). However, this does not fully account for the probability of stochastic genetic collision, as sampling in China was non-random and the genetic variant in question may be more frequent and introductions from China into Washington State may be more likely to occur from a subset of individuals within China. We additionally analyzed 293 Washington State viruses from the outbreak clade (excluding WA1) in a coalescent analysis to estimate temporal patterns. Here, we assume an evolutionary rate of ∼0.8 × 10^−3^ substitutions per site per year, consistent with rates for SARS-CoV-2 across the entire pandemic phylogeny. This analysis uses the degree and pattern of genetic diversity of sampled genomes to estimate the date of a common ancestor and exponential growth rate of the virus population. Applying it to these data gives a median estimate for the date of the clade’s common ancestor at 1 February 2020 with a 95% Bayesian credible interval of between 18 January and 9 February 2020 (**Fig. 1B**). This is consistent with either hypothesis (a) or (b) above. We additionally calculate a rate of exponential growth from the coalescent analysis for this clade, finding a median doubling time of 3.4 days with a 95% Bayesian credible interval of between 2.4 and 5.1 days (**Fig. 1B**).

In addition to the 293 viruses sampled from Washington State falling into the WA1 outbreak clade, we observe that seven viruses sampled from the Grand Princess cruise ship in late February and early Mar 2020 all group into the same outbreak clade (**Fig. 1A**). The genetic relationship among these viruses is consistent with a single introduction onto the Grand Princess cruise ship of the basal outbreak variant, possessing C17747T and A17858G, and subsequent transmission and evolution on the ship. With this phylogenetic structure, it is impossible to confidently determine whether the common ancestor of the outbreak clade is in Washington State, is on the Grand Princess, or was transmitted to both from an unsampled source. However, given the relative sizes of the Washington State outbreak and the Grand Princess outbreak, we believe a Washington State to Grand Princess transmission to be more likely, occurring subsequent to the introduction of the WA1 outbreak clade into Washington State.

In addition, we observed 53 SARS-CoV-2 genomes from Washington State that fall outside this primary outbreak clade and into multiple separate clusters (**Fig. 2**). Many of these separate clusters consist of single viruses and appear to be recent introductions not yet related to large clusters of local transmission. There is a second clade with 38 viruses that represents 11% of Washington State viruses. This clade is closely related to viruses from the European outbreak and likely represents a second introduction occurring at sometime in February 2020.

**Figure 2.**
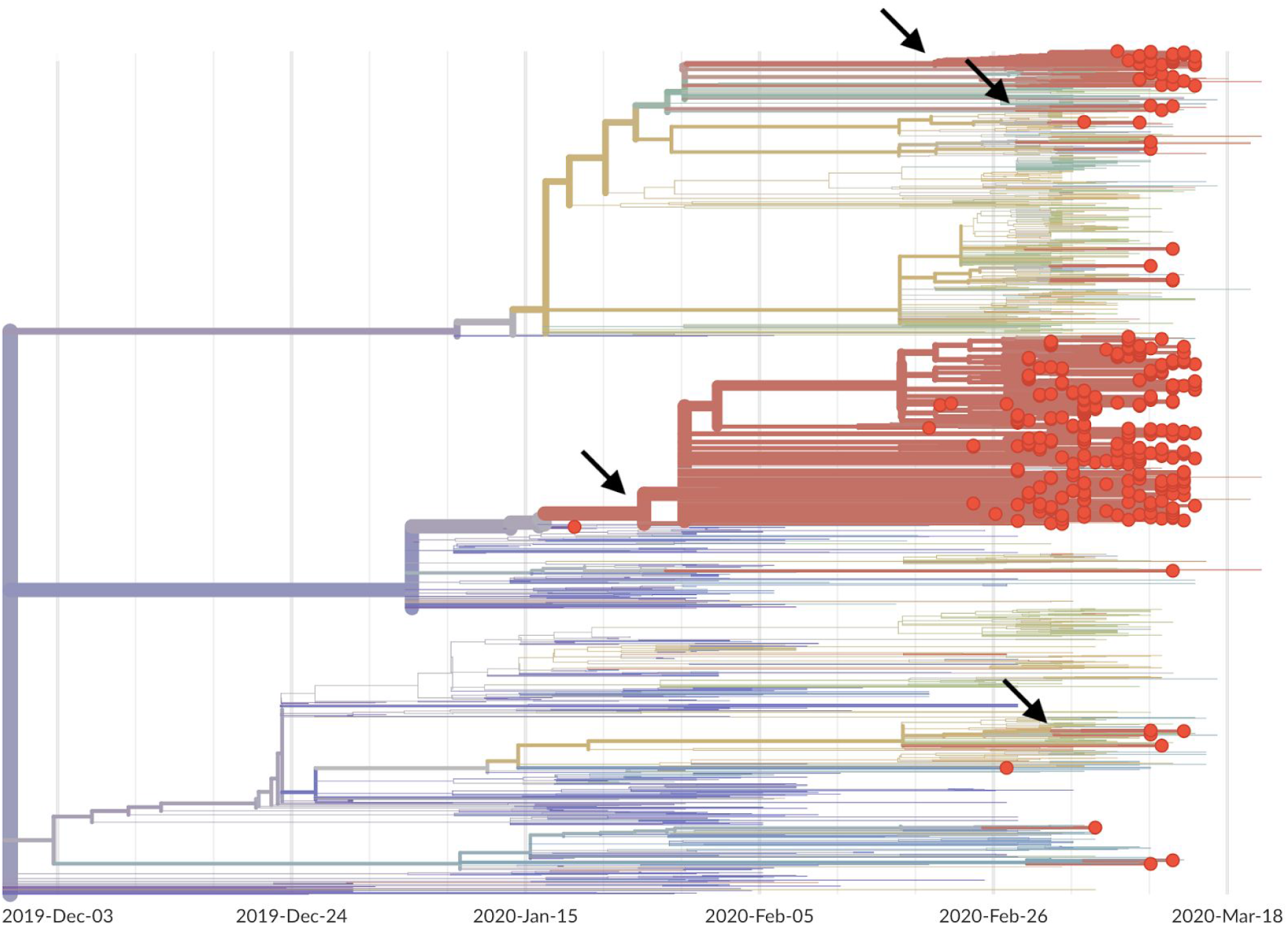
Phylogeny of 346 SARS-CoV-2 viruses collected from Washington State (red circles) on a background of globally collected viruses. Clustering of related viruses indicates community transmission after an introduction event. In addition to the estimated January introduction of the large outbreak clade we see later introduction events (marked by arrows) resulting in smaller community outbreaks.

Assuming a putative 15 January 2020 introduction based on the phylogenetic evidence, we simulated forward in a stochastic epidemiological transmission model to investigate the expected size of outbreak given this date of introduction. Based on epidemiological literature *(15 – 17)*, we chose a model with basic reproduction number *(R*0) of 3.2 corresponding to a mean doubling time of 6.1 days with 90% uncertainty interval of 5.1 to 8.2 days. Under this model, the median prevalence of active infections descended from the presumptive index case as of 1 March 2020 was 310 (90% uncertainty interval 50, 960), and with a total incidence to that date of 400 (90% uncertainty interval 80, 1300) infections (**Fig. 3**). As we have not yet been able to quantify the impact of social distancing policies since 5 March 2020, we estimate an upper bound of 1600 (90% uncertainty interval 250, 5100) active infections in this transmission chain by 15 March 2020. Although this simulation lacks explicit mobility, we expect the majority of downstream infections to be geographically localized. This model assumes a constant transmission rate; mitigation measures enacted during early March may have decreased transmission rate.

**Figure 3.**
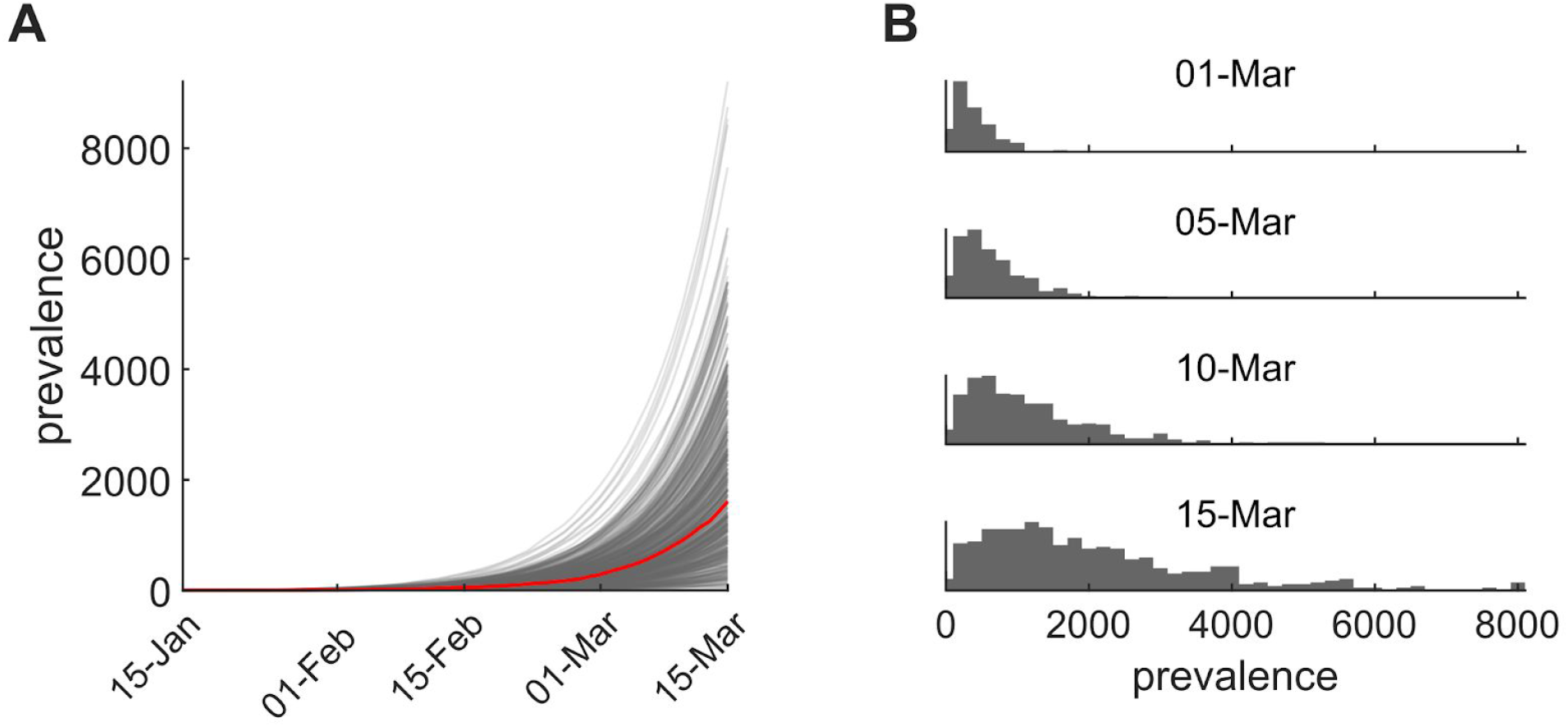
Inferences from the transmission model. (A) 500 prevalence trajectories showing the total number of people infected (presymptomatic/exposed and infectious) through time (median trajectory, red). Assumed start date is January 15 and simulations run through 15 March and do not account for impacts of social distancing policies since 5 March. (B) Distribution of simulated prevalences from 1 March 2020 to 15 March 2020.

In January and February, 2020, screening for SARS-CoV-2 in the United States was directed at travelers with fever, cough and shortness of breath, with the geographic areas increasing as new outbreaks were identified, but also specifying travel to China up until 24 February 2020 *(18, 19)*. Our analysis suggests that a single clade of SARS-CoV-2 had likely been circulating in the Seattle area for 4–6 weeks by the time the virus was first detected in a non-traveler on 28 Feb 2020. By then, variants within this clade constituted the majority of confirmed infections in the region (293 of 346;85%). Several factors could have contributed to the delayed detection of presumptive community spread, including limited testing among non-travelers or the presence of asymptomatic or mild illnesses. Genetic evidence suggests that this cluster may descend from an initial introduction in mid-January with the WA1 travel case, but other origin scenarios are also possible.

We demonstrate that SARS-CoV-2 was circulating in Washington State for 4-6 weeks before the first community-acquired case was detected on 28 February, 2020. Refining the time and geographic origin of the introduction into Washington State will require a combination of earlier samples and samples from other geographic locations, including from elsewhere in the United States and from China. It is possible that the Washington State outbreak originated from the WA1 introduction or from a separate introduction directly from China into Washington State, or from an introduction into Washington State from elsewhere in the United States. Given its size, the ongoing outbreak in New York City could also have resulted in early introduction(s) and cryptic community transmission. As of this date, the relative lack of genomic data from New York City limits what can be inferred about transmission there and how that relates to Washington State.

Our results highlight the critical need for widespread surveillance for community transmission of SARS-CoV-2 throughout the United States and the rest of the world even after the current pandemic is brought under control. The broad spectrum of disease severity *(20)* makes surveillance challenging *(21)*. The combination of traditional public health surveillance and genomic epidemiology can provide actionable insights, as happened in this instance: upon sequencing the initial community case on 29 February 2020, results were immediately shared via Twitter *(22)*, resulting in rapid rollout of social distancing policies as Seattle and Washington State came to grips with the extent of existing COVID-19 spread. From 29 February onwards, new genomic data was immediately posted to the GISAID EpiCoV sequence database *(23, 24)* and analyzed alongside other public SARS-CoV-2 genomes via the Nextstrain online platform *(25)* to provide immediate and public situational awareness. We see the combination of community surveillance, genomic analysis and public real-time sharing of results as empowering new systems for infectious disease surveillance.

## Ethics Approval

Sequencing and analysis of samples from the Seattle Flu Study was approved by the institutional review board at the University of Washington (protocol STUDY00006181). Informed consent was obtained for all community participant samples and survey data. Informed consent for residual sample and clinical data collection was waived. For UW Virology Lab, use of residual clinical specimens was approved by the institutional review board at the University of Washington (protocol STUDY00000408) with a waiver of informed consent.

## Disclaimer

This manuscript represents the opinions of the authors and does not necessarily reflect the position of the U.S. Centers for Disease Control and Prevention.

## Data Availability

Consensus SARS-CoV-2 genome sequences were deposited into GISAID immediately on generation. They have been submitted to Genbank as well (accession pending). All data used in the paper is also available here: https://github.com/blab/ncov-cryptic-transmission.

## Acknowledgments

We gratefully acknowledge the authors, originating and submitting laboratories of the sequences from GISAID’s EpiFlu™ Database on which this research is based. A full acknowledgements table is available as supplemental information. We have tried our best to avoid any direct analysis of genomic data not submitted as part of this paper and use this genomic data as background. We thank Joe Felsenstein and Chris Spitters for helpful discussion.

The Seattle Flu Study is run through the Brotman Baty Institute for Precision Medicine and funded by Gates Ventures, the private office of Bill Gates. The funder was not involved in the design of the study and does not have any ownership over the management and conduct of the study, the data, or the rights to publish. JS is an Investigator of the Howard Hughes Medical Institute. TB is a Pew Biomedical Scholar and is supported by NIH R35 GM119774-01. EBH and RAN are supported by University of Basel core funding. Sequencing analyses of SARS-CoV-2 genomes from California was supported by an NIH grant R33-AI129455 and the Charles and Helen Schwab Foundation to CYC, and an NIH grant K08-CA230156 and the Burroughs-Wellcome CAMS Award to WG.

## Seattle Flu Study Investigators

### Principal Investigators

Helen Y. Chu^1,7^, Michael Boeckh^1,2,7^, Janet A. Englund^3,7^, Michael Famulare^4^, Barry R. Lutz^5,7^, Deborah A. Nickerson^6,7^, Mark J. Rieder^7^, Lea M. Starita^6,7^, Matthew Thompson^9^, Jay Shendure^6,7,8^, and Trevor Bedford^2,6,7^

### Co-Investigators

Amanda Adler^3^, Elisabeth Brandstetter^1^, Shari Cho^6,7^, Chris D. Frazar^6^, Danielle Giroux^6^, Peter D. Han^6,7^, James Hadfield^1^, Shichu Huang^5^, Michael L. Jackson^10^, Anahita Kiavand^6,7^, Louise E. Kimball^2^, Kirsten Lacombe^3^, Jennifer Logue^1^, Victoria Lyon^1^, Kira L. Newman^1^, Matthew Richardson^6,7^, Thomas R. Sibley^2^, Monica L. Zigman Suchsland^1^, Melissa Truong^6,7^ and Caitlin

R. Wolf^1^

Affiliations:

1 Department of Medicine, University of Washington

2 Vaccine and Infectious Disease Division, Fred Hutchinson Cancer Research Center

3 Seattle Children’s Research Institute

4 Institute for Disease Modeling

5 Department of Bioengineering, University of Washington

6 Department of Genome Sciences, University of Washington

7 Brotman Baty Institute For Precision Medicine

8 Howard Hughes Medical Institute

9 Department of Family Medicine, University of Washington

10 Kaiser Permanente Washington Health Research Institute

## Materials and Methods

### Specimen collection

Specimens analyzed in this manuscript were obtained in a collaboration between the Washington State Department of Health, the Seattle Flu Study and the University of Washington Laboratory Medicine Department of Virology (UW Virology).

Specimens were collected by the Washington State Department of Health following CDC criteria. Specimens from UW Virology were obtained as part of clinical testing for SARS-CoV-2. Nasopharyngeal/oropharyngeal swabs were received from local healthcare providers and from the Washington State Department of Health to perform qualitative detection of SARS-CoV-2 RNA by a one-step real-time RT-PCR assay. Sequencing was performed on all samples with a positive or inconclusive RT-PCR assay result. The Seattle Flu Study (SFS) was established in the 2018-2019 flu season to combine clinical and innovative community sampling methods to measure how influenza, RSV, and other respiratory pathogens enter and circulate with the Seattle metropolitan region *(26)*. Samples screened for COVID-19 were collected as part of routine clinical testing, and residual samples were utilized for this study. Samples were additionally collected as part of prospective community enrollment of individuals with acute respiratory illness.

### Diagnostics and sequencing

Extracted nucleic acids (Magna Pure, 96 Roche) were screened for SARS-CoV-2 in a multiplexed Taqman assay with primer/probe sets targeting SARS-CoV-2 Orf1B (FAM) and human RNaseP (VIC) in duplicate (Life Technologies assay ID APGZJKF and A30064) in 384 well plates. Samples that are positive for SARS-CoV-2 are retested using the CDC-designed rtRT-PCR assays acquired directly from IDT (2019 nCoV Kit lot# 0000500389, 2019-nCoV_N positive control lot# 0000500326) according to CDC instructions with the exception that a 384-well plate and ViiA7 thermocycler was used.

SARS-CoV-2 genome sequencing was conducted using a metagenomic approach. RNA from positive specimens was converted to cDNA using random hexamers and reverse transcriptase (Superscript IV, Thermo) and a sequencing library was constructed using the Illumina TruSeq RNA Library Prep for Enrichment kit (Illumina). The library was sequenced on a MiSeq instrument using a V2 300 kit (Illumina). The resulting reads were assembled against the SARS-CoV-2 reference genome Wuhan-Hu-1/2019 (Genbank accession MN908947) using the bioinformatics pipeline https://github.com/seattleflu/assembly. Consensus sequences were deposited to Genbank (accessions pending) and GISAID.

For UW Virology samples, sequencing was performed as described previously *(27)*. Libraries were sequenced on Illumina MiSeq or NextSeq instruments using 1×185 or 1×75 runs respectively. Consensus sequences were assembled using a custom bioinformatics pipeline (https://github.com/proychou/hCoV19) adapted for SARS-CoV-2 from previous work *(28, 29)*. Briefly raw reads were trimmed to remove adapters and low quality regions using BBDuk and a k-mer based filter was used to pull out reads matching the reference sequence NC_045512. Filtered reads were *de novo* assembled using SPAdes *(30)* and contigs were ordered against the reference using BWA-MEM *(31)*. Gaps were filled by remapping reads against the assembled scaffold and a consensus sequence was called from this alignment using a custom script in R/Bioconductor. Consensus sequences were annotated using Prokka *(32)* and deposited to Genbank (accessions pending), GISAID, and NCBI SRA (Bioproject PRJNA610428).

We examined date of sample collection and other available metadata to remove possible duplicates from sequencing. The final dataset of 346 SARS-CoV-2 genomes from Washington State represent a convenience sample from the underlying outbreak. All Washington State sequences used in the paper are available here https://github.com/blab/ncov-cryptic-transmission.

### Phylogenetics

SARS-CoV-2 genomes from the global COVID-19 pandemic were downloaded from GISAID *(23, 24)* and processed using the Nextstrain *(25)* bioinformatics pipeline *Augur* to align genomes via MAFFT v7.4 *(33)*, build maximum likelihood phylogeny via IQTREE v1.6 *(34)* and reconstruct nucleotide and amino acid changes on the ML tree. This bioinformatic processing pipeline is fully documented and reproducible at https://github.com/nextstrain/ncov. The resulting tree was visualized in the Nextstrain web application *Auspice* to view resulting inferences.

Additionally, 293 SARS-CoV-2 aligned genomes from the WA1 outbreak clade were analyzed in BEAST *(35)* to estimate time of common ancestor and rate of epidemic growth. This analysis used an exponential growth coalescent model in which effective population size and rate of exponential growth are estimated. We assumed a HKY85 nucleotide substitution model *(36)* with gamma distributed rate variation and a strict molecular clock with a mean of 0.8 × 10^−3^ substitutions per site per year. Full analysis details, including BEAST XML, are available at https://github.com/blab/ncov-cryptic-transmission.

### Dynamical modeling

To model plausible ranges for the cumulative incidence and current prevalence following the introduction of SARS-CoV-2 into Snohomish County, WA, from the presumptive index case described in ref. *(12)*, we used a stochastic susceptible-exposed-infectious-recovered (SEIR) model with the following assumptions. The exposed (latent) period prior to the onset of viral shedding is normally-distributed with a mean of 4 days and standard deviation of 1 day; this is one day shorter than the 5 day consensus estimate of the incubation period prior to symptom onset (MIDAS-network) to acknowledge reports of pre-symptomatic shedding. The infectious period is normally distributed with mean 8 days and standard deviation 2 days, based on measured upper-respiratory viral shedding after symptom onset *(37)*. We drew transmission events from a truncated normal distribution to approximately reproduce the negative binomial transmission dynamics with transmission heterogeneity parameter *k*=0.54 from ref. *(38)*. We chose a truncated normal to reproduce the general overdispersion associated with SARS-CoV-2 transmission but with reduced probability of very large events involving more than twenty transmissions from a single person. For initial conditions, we assumed that the introduction into Snohomish County began on 15 Jan, 2020, coincident with the return of the presumptive index case to western Washington from Wuhan, China and with their symptom onset date *(12)*. The model code and scripts to run it are available at https://github.com/blab/ncov-cryptic-transmission.

## Supplemental Figures

**Supplementary Fig. 1.**
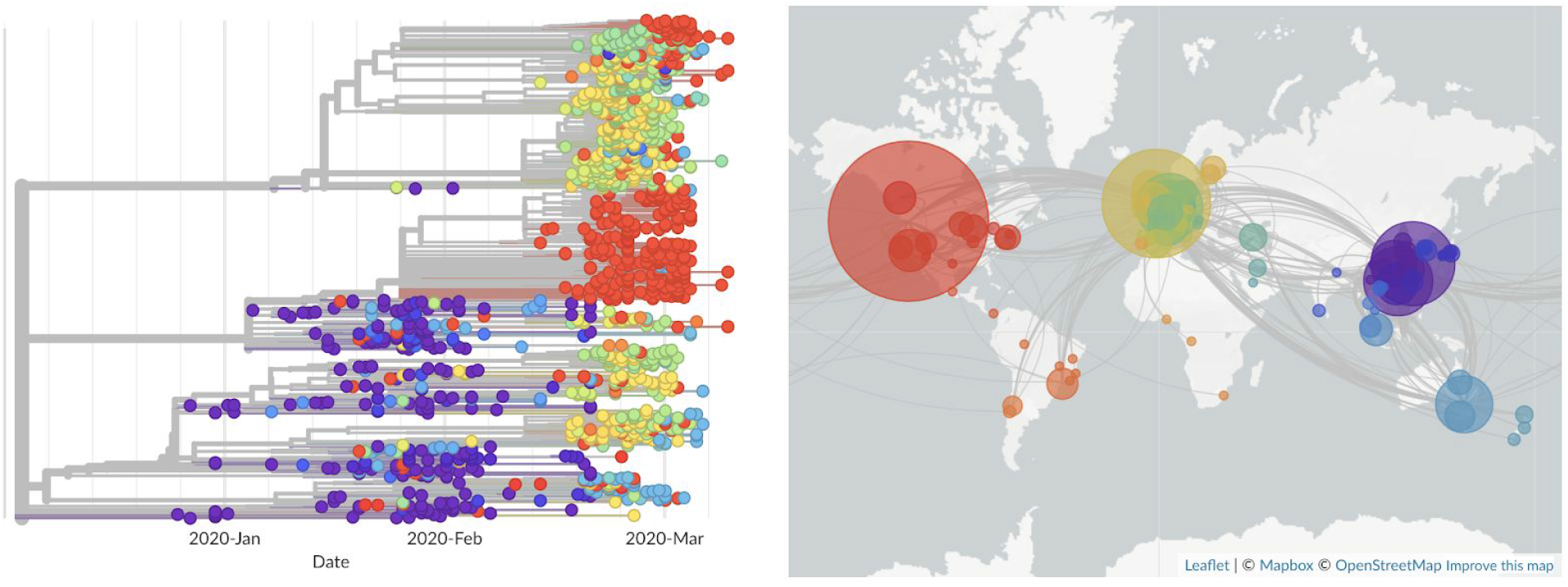
Phylogeny of 1442 SARS-CoV-2 viruses collected between December 2019 and March 2020 colored by country of sampling as shown in the map on the right. Viruses from China are shown in purple, viruses from Southeast Asia and Oceania in blue, viruses from Europe in yellow/green, viruses from South America in orange, and viruses from North America in red. Virus genome data shared through GISAID and phylogeny reconstructed by Nextstrain.

## References

1. World Health Organization, WHO Director-General’s opening remarks at the media briefing on COVID-19 - 11 March 2020 (2020), (available at https://www.who.int/dg/speeches/detail/who-director-general-s-opening-remarks-at-the-media-briefing-on-covid-19--11-march-2020).

2. A. E. Gorbalenya, Severe acute respiratory syndrome-related coronavirus--The species and its viruses, a statement of the Coronavirus Study Group. BioRxiv (2020) (available at https://www.biorxiv.org/content/10.1101/2020.02.07.937862v1.abstract).

3. S. Jiang, Z. Shi, Y. Shu, J. Song, G. F. Gao, W. Tan, D. Guo, A distinct name is needed for the new coronavirus. Lancet (2020), doi:10.1016/S0140-6736(20)30419-0.

4. Rambaut A, Phylogenetic analysis of nCoV-2019 genomes. Virological, (available at http://virological.org/t/phylodynamic-analysis-176-genomes-6-mar-2020/356).

5. World Health Organization, “Coronavirus disease 2019 (COVID-19) Situation report 55” (2020, March 15), (available at https://www.who.int/emergencies/diseases/novel-coronavirus-2019/situation-reports).

6. F. Schlosser, B. F. Maier, O. Baranov, D. Brockmann, C. Jongen, A. Zachariae, A. Rose, Coronavirus COVID-19 Global Risk Assessment. Event Horizon - COVID-19, (available at http://rocs.hu-berlin.de/corona/).

7. World Health Organization, “Coronavirus disease 2019 (COVID-19) Situation report 38” (2020, Feb 27), (available at https://www.who.int/emergencies/diseases/novel-coronavirus-2019/situation-reports).

8. Kari Bray, Coronavirus update: Addressing questions about a presumptive positive case in an adolescent. Snohomish Health District Public Health Essentials (2020), (available at https://www.snohd.org/Blog.aspx?IID=13).

9. Washington State Department of Health, 2019 Novel Coronavirus (COVID-19) in Washington (2020), (available at https://www.doh.wa.gov/Emergencies/Coronavirus).

10. K. G. Andersen, A. Rambaut, W. I. Lipkin, E. C. Holmes, R. F. Garry, The proximal origin of SARS-CoV-2. ARTIC Network. 17 (2020).

11. N. Imai, I. Dorigatti, A. Cori, C. Donnelly, S. Riley, N. M. Ferguson, “Report 2: Estimating the potential total number of novel Coronavirus cases in Wuhan City, China” (MRC Centre for Global Infectious Disease Analysis, 2020).

12. M. L. Holshue, C. DeBolt, S. Lindquist, K. H. Lofy, J. Wiesman, H. Bruce, C. Spitters, K. Ericson, S. Wilkerson, A. Tural, G. Diaz, A. Cohn, L. Fox, A. Patel, S. I. Gerber, L. Kim, S. Tong, X. Lu, S. Lindstrom, M. A. Pallansch, W. C. Weldon, H. M. Biggs, T. M. Uyeki, S. K. Pillai, Washington State 2019-nCoV Case Investigation Team, First Case of 2019 Novel Coronavirus in the United States. N. Engl. J. Med. (2020), doi:10.1056/NEJMoa2001191.

13. Q. Li, X. Guan, P. Wu, X. Wang, L. Zhou, Y. Tong, R. Ren, K. S. M. Leung, E. H. Y. Lau, J. Y. Wong, X. Xing, N. Xiang, Y. Wu, C. Li, Q. Chen, D. Li, T. Liu, J. Zhao, M. Liu, W. Tu, C. Chen, L. Jin, R. Yang, Q. Wang, S. Zhou, R. Wang, H. Liu, Y. Luo, Y. Liu, G. Shao, H. Li, Z. Tao, Y. Yang, Z. Deng, B. Liu, Z. Ma, Y. Zhang, G. Shi, T. T. Y. Lam, J. T. Wu, G. F. Gao, B. J. Cowling, B. Yang, G. M. Leung, Z. Feng, Early Transmission Dynamics in Wuhan, China, of Novel Coronavirus–Infected Pneumonia. N. Engl. J. Med. (2020), doi:10.1056/NEJMoa2001316.

14. H. Nishiura, N. M. Linton, A. R. Akhmetzhanov, Serial interval of novel coronavirus (COVID-19) infections. Int. J. Infect. Dis. (2020), doi:10.1016/j.ijid.2020.02.060.

15. J. T. Wu, K. Leung, G. M. Leung, Nowcasting and forecasting the potential domestic and international spread of the 2019-nCoV outbreak originating in Wuhan, China: a modelling study. Lancet (2020), doi:10.1016/S0140-6736(20)30260-9.

16. M. Chinazzi, J. T. Davis, M. Ajelli, C. Gioannini, M. Litvinova, S. Merler, A. Pastore y Piontti, L. Rossi, K. Sun, C. Viboud, X. Xiong, H. Yu, M. E. Halloran, I. M. Longini Jr., A. Vespignani, The effect of travel restrictions on the spread of the 2019 novel coronavirus (2019-nCoV) outbreak. Epidemiology (2020), doi:10.1101/2020.02.09.20021261.

17. S. Sanche, Y. T. Lin, C. Xu, E. Romero-Severson, N. Hengartner, R. Ke, The Novel Coronavirus, 2019-nCoV, is Highly Contagious and More Infectious Than Initially Estimated. Epidemiology (2020),, doi:10.1101/2020.02.07.20021154.

18. Update and Interim Guidance on Outbreak of 2019 Novel Coronavirus (2019-nCoV). CDC Health Alert Network (2020), (available at https://emergency.cdc.gov/han/han00427.asp).

19. US Centers for Disease Control and Prevention, Criteria to Guide Evaluation of Persons Under Investigation (PUI) for 2019-nCoV (2020), (available at https://web.archive.org/web/20200222215422/https://www.cdc.gov/coronavirus/2019-ncov/hcp/clinical-criteria.html).

20. W.-J. Guan, Z.-Y. Ni, Y. Hu, W.-H. Liang, C.-Q. Ou, J.-X. He, L. Liu, H. Shan, C.-L. Lei, D. S. C. Hui, B. Du, L.-J. Li, G. Zeng, K.-Y. Yuen, R.-C. Chen, C.-L. Tang, T. Wang, P.-Y. Chen, J. Xiang, S.-Y. Li, J.-L. Wang, Z.-J. Liang, Y.-X. Peng, L. Wei, Y. Liu, Y.-H. Hu, P. Peng, J.-M. Wang, J.-Y. Liu, Z. Chen, G. Li, Z.-J. Zheng, S.-Q. Qiu, J. Luo, C.-J. Ye, S.-Y. Zhu, N.-S. Zhong, China Medical Treatment Expert Group for Covid-19, Clinical Characteristics of Coronavirus Disease 2019 in China. N. Engl. J. Med. (2020), doi:10.1056/NEJMoa2002032.

21. R. Li, S. Pei, B. Chen, Y. Song, T. Zhang, W. Yang, J. Shaman, Substantial undocumented infection facilitates the rapid dissemination of novel coronavirus (SARS-CoV2). Science (2020), doi:10.1126/science.abb3221.

22. T. Bedford, The team at the @seattleflustudy have sequenced the genome the #COVID19 community case reported yesterday from Snohomish County, WA, and have posted the sequence publicly to http://gisaid.org. There are some enormous implications here. Twitter (2020), (available at https://twitter.com/trvrb/status/1233970271318503426).

23. Y. Shu, J. McCauley, GISAID: Global initiative on sharing all influenza data - from vision to reality. Euro Surveill. 22 (2017), doi:10.2807/1560-7917.ES.2017.22.13.30494.

24. S. Elbe, G. Buckland-Merrett, Data, disease and diplomacy: GISAID’s innovative contribution to global health. Glob Chall. 1, 33–46 (2017).

25. J. Hadfield, C. Megill, S. M. Bell, J. Huddleston, B. Potter, C. Callender, P. Sagulenko, T. Bedford, R. A. Neher, Nextstrain: real-time tracking of pathogen evolution. Bioinformatics. 34, 4121–4123 (2018).

26. H. Y. Chu, M. Boeckh, J. A. Englund, M. Famulare, B. R. Lutz, D. A. Nickerson, M. Rieder, L. Starita, M. Thompson, J. Shendure, T. Bedford, A. Adler, E. Brandstetter, J. Bosua, C. D. Frazar, P. D. Han, R. Gulati, J. Hadfield, S. Huang, M. L. Jackson, A. Kiavand, L. E. Kimball, K. Lacombe, J. Logue, V. Lyon, T. R. Sibley, M. L. Zigman Suchsland, C. R. Wolf, LB 21. The Seattle Flu Study: A Community-Based Study of Influenza. Open Forum Infect Dis. 6, S1002–S1002 (2019).

27. A. L. Greninger, D. M. Zerr, X. Qin, A. L. Adler, R. Sampoleo, J. M. Kuypers, J. A. Englund, K. R. Jerome, Rapid Metagenomic Next-Generation Sequencing during an Investigation of Hospital-Acquired Human Parainfluenza Virus 3 Infections. J. Clin. Microbiol. 55, 177–182 (2017).

28. A. L. Greninger, G. M. Knudsen, P. Roychoudhury, D. J. Hanson, R. H. Sedlak, H. Xie, J. Guan, T. Nguyen, V. Peddu, M. Boeckh, M.-L. Huang, L. Cook, D. P. Depledge, D. M. Zerr, D. M. Koelle, S. Gantt, T. Yoshikawa, M. Caserta, J. A. Hill, K. R. Jerome, Comparative genomic, transcriptomic, and proteomic reannotation of human herpesvirus 6. BMC Genomics. 19, 204 (2018).

29. A. L. Greninger, P. Roychoudhury, H. Xie, A. Casto, A. Cent, G. Pepper, D. M. Koelle, M.-L. Huang, A. Wald, C. Johnston, K. R. Jerome, Ultrasensitive Capture of Human Herpes Simplex Virus Genomes Directly from Clinical Samples Reveals Extraordinarily Limited Evolution in Cell Culture. mSphere. 3 (2018), doi:10.1128/mSphereDirect.00283-18.

30. A. Bankevich, S. Nurk, D. Antipov, A. A. Gurevich, M. Dvorkin, A. S. Kulikov, V. M. Lesin, S. Nikolenko, S. Pham, A. D. Prjibelski, A. V. Pyshkin, A. V. Sirotkin, N. Vyahhi, G. Tesler, M. A. Alekseyev, P. A. Pevzner, SPAdes: a new genome assembly algorithm and its applications to single-cell sequencing. J. Comput. Biol. 19, 455–477 (2012).

31. M. Vasimuddin, S. Misra, H. Li, S. Aluru, Efficient Architecture-Aware Acceleration of BWA-MEM for Multicore Systems. 2019 IEEE International Parallel and Distributed Processing Symposium (IPDPS) (2019),, doi:10.1109/ipdps.2019.00041.

32. T. Seemann, Prokka: rapid prokaryotic genome annotation. Bioinformatics. 30, 2068–2069 (2014).

33. K. Katoh, D. M. Standley, MAFFT multiple sequence alignment software version 7: improvements in performance and usability. Mol. Biol. Evol. 30, 772–780 (2013).

34. L.-T. Nguyen, H. A. Schmidt, A. von Haeseler, B. Q. Minh, IQ-TREE: a fast and effective stochastic algorithm for estimating maximum-likelihood phylogenies. Mol. Biol. Evol. 32, 268–274 (2015).

35. M. A. Suchard, P. Lemey, G. Baele, D. L. Ayres, A. J. Drummond, A. Rambaut, Bayesian phylogenetic and phylodynamic data integration using BEAST 1.10. Virus Evol. 4, vey016 (2018).

36. M. Hasegawa, H. Kishino, T. Yano, Dating of the human-ape splitting by a molecular clock of mitochondrial DNA. J. Mol. Evol. 22, 160–174 (1985).

37. L. Zou, F. Ruan, M. Huang, L. Liang, H. Huang, Z. Hong, J. Yu, M. Kang, Y. Song, J. Xia, Q. Guo, T. Song, J. He, H.-L. Yen, M. Peiris, J. Wu, SARS-CoV-2 Viral Load in Upper Respiratory Specimens of Infected Patients. N. Engl. J. Med. (2020), doi:10.1056/NEJMc2001737.

38. J. Riou, C. L. Althaus, Pattern of early human-to-human transmission of Wuhan 2019-nCoV. bioRxiv (2020), p. 2020.01.23.917351.

